# Reproducibility of 3D MRSI for imaging human brain glucose metabolism using direct (^2^H) and indirect (^1^H) detection of deuterium labeled compounds at 7T and clinical 3T

**DOI:** 10.1101/2023.04.17.23288672

**Authors:** Fabian Niess, Bernhard Strasser, Lukas Hingerl, Eva Niess, Stanislav Motyka, Gilbert Hangel, Martin Krššák, Stephan Gruber, Benjamin Spurny-Dworak, Siegfried Trattnig, Thomas Scherer, Rupert Lanzenberger, Wolfgang Bogner

## Abstract

**Introduction:** Deuterium metabolic imaging (DMI) and quantitative exchange label turnover (QELT) are novel MR spectroscopy techniques for non-invasive imaging of human brain glucose and neurotransmitter metabolism with high clinical potential. Following oral or intravenous administration of non-ionizing [6,6’-^2^H_2_]-glucose, its uptake and synthesis of downstream metabolites can be mapped via direct or indirect detection of deuterium resonances using ^2^H MRSI (DMI) and ^1^H MRSI (QELT), respectively.

The purpose of this study was to compare the dynamics of spatially resolved brain glucose metabolism, i.e., estimated concentration enrichment of deuterium labeled Glx (glutamate+glutamine) and Glc (glucose) acquired repeatedly in the same cohort of subjects using DMI at 7T and QELT at clinical 3T.

**Methods:** Five volunteers (4m/1f) were scanned in repeated sessions for 60 min after overnight fasting and 0.8g/kg oral [6,6’-^2^H_2_]-glucose administration using time-resolved 3D ^2^H FID-MRSI with elliptical phase encoding at 7T and 3D ^1^H FID-MRSI with a non-Cartesian concentric ring trajectory readout at clinical 3T.

**Results:** One hour after oral tracer administration regionally averaged deuterium labeled Glx_4_ concentrations and the dynamics were not significantly different over all participants between 7T ^2^H DMI and 3T ^1^H QELT data for GM (1.29±0.15 vs. 1.38±0.26 mM, p=0.65 & 21±3 vs. 26±3 µM/min, p=0.22) and WM (1.10±0.13 vs. 0.91±0.24 mM, p=0.34 & 19±2 vs. 17±3 µM/min, p=0.48). Also, the observed time constants of dynamic Glc_6_ data in GM (24±14 vs. 19±7 min, p=0.65) and WM (28±19 vs. 18±9 min, p=0.43) dominated regions showed no significant differences.

Between individual ^2^H and ^1^H data points a weak to moderate negative correlation was observed for Glx_4_ concentrations in GM (r=-0.52, p<0.001), and WM (r=-0.3, p<0.001) dominated regions, while a strong negative correlation was observed for Glc_6_ data GM (r=- 0.61, p<0.001) and WM (r=-0.70, p<0.001).

**Conclusion:** This study demonstrates that indirect detection of deuterium labeled compounds using ^1^H QELT MRSI at widely available clinical 3T without additional hardware is able to reproduce absolute concentration estimates of downstream glucose metabolites and the dynamics of glucose uptake compared to ^2^H DMI data acquired at 7T. This suggests significant potential for widespread application in clinical settings especially in environments with limited access to ultra-high field scanners and dedicated RF hardware.

## Introduction

Glucose is the main energy source in the mammalian brain and its metabolism provides fuel for physiological brain function and cellular maintenance via adenosine triphosphate (ATP) synthesis. Moreover, oxidative glucose metabolism generates precursors required for neurotransmitter biosynthesis, e.g., glutamate (1, 2). Several common pathologies are characterized by alterations in brain glucose uptake compared to healthy tissue as a result of impaired glucose metabolism, e.g., Alzheimer’s disease (3), depression and schizophrenia (4), tissue ischemia and cancer (5).

[^18^F]-Fluorodeoxyglucose ([^18^F]-FDG) positron emission tomography (PET) is the current clinical gold standard to image tissue specific glucose uptake. However, it does not provide information about downstream metabolites, e.g., oxidative neurotransmitter synthesis of glutamate or glycolytic lactate production due to glucose trapping of the tracer and is an invasive technique, since FDG is radioactive (6, 7).

Deuterium metabolic imaging (DMI) is as novel technique for the non-invasive mapping of brain glucose metabolism using time resolved ^2^H Magnetic Resonance Spectroscopic Imaging (MRSI) after oral or intravenous administration of harmless and safe deuterium labeled glucose ([6,6’]-^2^H-Glc) as tracer (8, 9). A simultaneous detection of glucose uptake and downstream metabolic products, such as glutamate, glutamine and lactate provide complementary information and allows for separating oxidative from non-oxidative metabolic pathways during glucose metabolism (10, 11). However, modifications of the MR scanner configuration and external frequency sources may be required to facilitate ^2^H DMI as transmission and reception of ^2^H Larmor frequency is not widely supported by all MR vendors and additional dedicated RF equipment is needed (12–14). Furthermore, the majority of human DMI applications was performed at ultra-high magnetic field strengths (≥4T).

Recently quantitative exchange label turnover (QELT) has been introduced, which indirectly detects accumulation of deuterium labeled metabolites using conventional time resolved ^1^H MRS/MRSI at ultra-high magnetic field strength (≥7T) (15–18) and clinical 3T (19). Due to an exchange of deuterium labeled and unlabeled molecules, a signal decrease of the resonance of the respective metabolite can be detected in conventional ^1^H MR spectra, similarly as performed using ^13^C labeled glucose (20, 21). In contrast to DMI, no additional hardware is required, such as dual-tuned dedicated RF coils or modifications of the MR scanner itself.

Additionally, an extended neurochemical profile is quantified as QELT simultaneously detects other resonances present in ^1^H MR spectra, which are not involved in glucose metabolism.

The aim of the study was to assess the reproducibility of the QELT method compared to the more common DMI approach for monitoring the dynamics of spatially resolved glucose uptake and oxidative Glx synthesis. Both direct (^2^H DMI) and indirect (^1^H QELT) deuterium detection methods were employed to acquire time-resolved 3D data from the same cohort of participants, repeatedly, at 7T and clinical 3T, respectively. The concentration estimates were quantified using internal referencing to evaluate and compare the performance of both methods.

## Methods

### Study protocol

Approved by the local ethics committee of the Medical University of Vienna, this study included two separate MRI protocols, i.e., time-resolved deuterium metabolic imaging (DMI) and quantitative exchange label turnover (QELT) conducted on separate days, 1-3 months apart, on an experimental 7T and clinical 3T MR systems, respectively. Before, during and after the initial DMI protocol, capillary puncture blood sampling was performed every 15 min over the course of 1.5 hours from the toe (accessible sampling site without moving the volunteer out of the MR scanner) using two identical standard strip glucometers (Verio, OneTouch) for cross checking.

### Volunteers

Five lean volunteers without history of neurological, psychiatric or metabolic diseases (4 male/ 1 female, BMI: 22±1 kg/m^2^, age: 33±5 years) were recruited and gave written informed consent to participate in this study. For both DMI and QELT MRI protocols, volunteers were scanned after overnight fasting and immediately after oral tracer administration using 0.8 g/kg body weight deuterium-labeled glucose ([6,6’]-2H-Glc ≥99% purity, Cambridge Isotopes) dissolved in 200 ml water. The tracer was consumed within one minute, immediately before volunteers were moved inside the scanner bore.

### 7T ^2^H DMI protocol

^2^H DMI data acquisition was supported by the vendor, without any modification of the scanner hardware. Data were acquired on a Siemens 7T (dot Plus) whole body MR system using a 1 channel transmit/ 32 channel receive ^1^H head coil (Nova Medical, Wilmington, MA, USA) and a dual-tuned (^2^H/^1^H) quadrature transmit-receiver birdcage coil (Stark Contrast MRI Coils Research, Germany) to acquire anatomical high-resolution ^1^H MR images and 3D ^2^H MR spectroscopic images, respectively. The DMI protocol includes initial preparation scans, i.e., unlocalized pulse-acquire B_1_ mapping (T_R_=1500 ms, T_E_=0.35 ms, 20 steps, U_Ref_=20- 440 V) to estimate the required reference voltage followed by 10 consecutive 3D FID MRSI scans using elliptical phase encoding with the following parameters: FOV: 200x200x175 mm^3^, matrix size: 16x16x14, nominal voxel volume: 1.95 ml, 2 averages, T_R_=290 ms, T_E_=1.5 ms, T_A_=6:37 min, BW=500 Hz, 128 samples, flip angle: 86° (for additional sequence details see Supplementary Table 1). After each 3D MRSI dataset, the frequency was updated to account for potential frequency drifts. High resolution T_1_-weighted 3D MP2RAGE images were acquired using both dual-tuned (_2_H/_1_H) birdcage head coil and 32 channel receiver head coils after the subject was repositioned using the following parameters: 1.1mm_3_ isotropic nominal voxel volume, FOV: 165x220x220 mm^3^, grid size: 144x192x192, T_R_=3930 ms, T_I1_=850 ms, T_I2_=3400 ms T_E_=3.28 ms, 3-fold GRAPPA accelerated and T_A_=4:29 min (using 32 channel head coil). Following co-registration of both 3D images, the latter were then used for tissue segmentation.

### 3T ^1^H QELT protocol

^1^H QELT data were acquired on a clinical routine Siemens 3T MR system (Prisma-Fit) using a 64-channel receiver head coil (Siemens Healthineers, Erlangen, Germany). The QELT protocol included an initial automated alignment localizer and EPI reference scans to set up the volumetric navigator sequence used for real-time motion correction (22). Following sequence preparation, 14 consecutive 3D ^1^H MRSI datasets were acquired over the course of ∼60 min using a previously developed 3D ^1^H FID MRSI sequence with slice-selective excitation and a fast concentric ring trajectory readout including automatic interleaved real-time motion-, shim- and frequency drift correction (23, 24). The following parameters were applied (19): 0.24 ml isotropic nominal voxel volume, FOV: 200x200x130 mm^3^, VOI: 200x200x55 mm^3^, grid size: 32x32x21, centered around the posterior cingulate region, T_E_=0.8 ms, T_R_ = 950 ms and T_A_=4:13 min (details see supplemental material table S2 for minimum reporting standards (25)). Within each T_R_ unsuppressed water reference signals (20 FID points) were acquired using identical readout trajectories to approximate the coil sensitivity for each channel followed by a conventional WET water suppression scheme (26). Following the QELT MRSI scan a high resolution T_1_ weighted 3D MPRAGE scan was performed for anatomical imaging and tissue segmentation with the following parameters: 1mm^3^ isotropic nominal voxel volume, FOV: 208x250x250 mm^3^, grid size: 208x256x256, T_R_=1800 ms, T_I_=900 ms, T_E_=2.27 ms, 3-fold GRAPPA accelerated and T_A_=2:38 min.

**Table 2:** LCModel spectral fit output of a single representative GM and WM voxel for selected ^2^H and ^1^H metabolites from the first (5 min) and last (60 min) time point after deuterium labeled glucose ingestion, acquired using 7T ^2^H DMI and 3T ^1^H QELT, respectively.

| Metabolite | Gray matter voxel |  | White matter voxel |  |
| --- | --- | --- | --- | --- |
|  | concentration [a.u.] (CRLB [%]) |  | concentration [a.u.] (CRLB [%]) |  |
|  | first time point<br>[a.u. (CRLB %)] | last time point<br>[a.u. (CRLB %)] | first time point<br>[a.u. (CRLB %)] | last time point<br>[a.u. (CRLB %)] |
| **7T $^2\text{H}$ DMI** |  |  |  |  |
| $^2\text{H}$ water | 4.68 (3%) | 5.38 (3%) | 4.29 (3%) | 5.13 (3%) |
| Glx<sub>4</sub> | 0.08 (154%) | 1.84 (9%) | 0.29 (46%) | 1.48 (10%) |
| Glc<sub>6</sub> | 0.26 (50%) | 1.58 (10%) | 0.18 (72%) | 1.54 (9%) |
| **3T $^1\text{H}$ QELT** |  |  |  |  |
| tCr | 9.40E5 (4%) | 8.92E5 (5%) | 7.80E5 (4%) | 7.09E5 (5%) |
| Glx<sub>4</sub> | 1.65E6 (7%) | 1.31E6 (10%) | 1.06E6 (10%) | 8.18E5 (14%) |
| Glc<sub>6</sub> | 5.54E5 (22%) | 3.79E5 (27%) | 5.69E5 (18%) | 3.06E5 (35%) |

Notes. – $^2\text{H}$ water = deuterated water, Glx<sub>4</sub> = combined glutamate+glutamine (4<sup>th</sup> carbon position resonance), Glc<sub>6</sub> = glucose beta anomer (6<sup>th</sup> carbon position resonance), tCr= total creatine

### Data Reconstruction

All data were reconstructed offline using a custom-built automated software pipeline (MATLAB R2021, bash, Python3.10).

The Cartesian phase-encoded ^2^H DMI MRSI data were Hamming filtered in all three spatial dimensions followed by a three dimensional Fourier transform.

For ^1^H QELT 3D MRSI data acquired using non-cartesian concentric ring trajectories the pipeline included in-plane convolutional re-gridding of the k-space (27), noise-decorrelation, channel-wise lipid decontamination (28) and coil combination (29) using weights determined by water unsuppressed pre-scans from each T_R_.

### Spectral Fitting/Metabolite Quantification

Spectral fitting was performed voxel-wise in the frequency domain using LCModel (v6.3) (30). Quantification results with Cramer-Rao Lower Bounds (CRLB) >20% were excluded from further analysis. For ^1^H: Glc_6_ and ^2^H: Glx_4_ and Glc_6_ results a CRLB threshold of 50% was used. No CRLB threshold was used for the first 3 time points of ^2^H fit results (first 20 min). Spectral fitting of ^2^H DMI MRSI data used a custom-built basis set including simulated (31, 32) ^2^H resonances of water, glucose (Glc) and combined Glx.

For spectral quantification of ^1^H QELT MRSI data a modified basis set was used featuring 17 neurochemical metabolites (i.e., creatine, phosphocreatine, myo-inositol, N-acetylaspartate, N-acetylaspartylglutamate, glutathione, glycerophosphocholine, phosphocholine, aspartate, glucose-alpha, glucose-beta, taurine, glutamate, glutamine, gamma-aminobutyric acid, lactate) and a measured macromolecular background (33). This work focuses only on few relevant metabolites, i.e., tCr, Glu_4_+Gln_4_ (Glx_4_), and Glc_6._

Deuterium-labeled glucose ([6,6’]-^2^H-Glc) features deuterium atoms on the 6^th^ carbon position and during metabolic utilization particular downstream metabolites incorporate deuterium at specific carbon positions only, e.g., oxidatively synthesized glutamate and glutamine labeling occurs at the 4^th^ carbon position in the brain. Therefore, Glc and Glx peaks in respective ^2^H MR spectra represent Glc_6_ (3.8 ppm) and Glx_4_ (2.3 ppm) resonances. While deuterium labeled resonances do not directly contribute to signals visible in ^1^H MR spectra, increasing levels of labeled Glx and Glc molecules eventually lead to a respective signal decrease of Glc_6_ and Glx_4_ resonances in the ^1^H spectrum, due to an exchange with unlabeled molecules, indirectly reflecting deuterium enrichment. Other ^1^H resonances of the same molecule (Glc_1-5_, and Glx_23_) remain stable. Therefore, using ^1^H MRS to reliably detect the signal decrease of a single molecular resonance only, labeled (Glu_4_=2.34 ppm, Gln_4_=2.44 ppm, Glc_6_=3.88 ppm) and combined unlabeled parts (Glu_23,_ Gln_23,_ Glc_1-5_: including Glu_2_=3,75 ppm, Glu_3_=2.10 ppm, Gln_2_=3.77 ppm, Gln_3_=2.13ppm, Glc_1_=4.63 ppm, Glc_2_=3.23 ppm, Glc_3_=3.47 ppm, Glc_4_=3.38 ppm, Glc_5_=3.45 ppm) of the respective metabolites were separated in the basis set to be fitted individually. To take J-coupling effects into account Glu_4_, Gln_4_ and Glc_6_ resonances were created by simulating Glu_23_, Gln_23_ or Glc_1-5_ (fully deuterated state: both protons are exchanged with deuterons) and subtraction from regular Glu, Gln, or Glc signals.

### Concentration estimation

For all time points of 7T DMI and 3T QELT data 3D metabolite maps were created. Glx_4_ (^2^H and ^1^H resonances) and Glc_6_ (^2^H resonance) concentrations were given in mM units, while Glc_6_ (^1^H resonance) maps were given as ratio to total creatine as to the best of our knowledge no relaxation times were reported for ^1^H glucose at 3T. Concentration estimates were calculated as presented in (34), using natural abundance deuterated ^2^H water (averaged over the first 3 time points) and ^1^H total creatine as internal reference, for 7T DMI and 3T QELT, respectively. Relaxation times and in vivo concentrations were assumed using literature values (9, 14, 35–37): ([^2^H: water_T1/T2_=350 ms/30 ms, Glx_T1/T2_=150 ms/40 ms, Glc_T1/T2_=67 ms/42 ms, ^2^H water concentration: 17.2 mM], [^1^H T_1_: tCr_GM/WM_= 1.46 s/1.24 s, Glx_GM/WM_= 1.27 s/1.24 s, T_2_: tCr_GM/WM_ = 201 ms/198 ms, Glx_GM/WM_= 134 ms/148 ms], tCr_GM_ = 7.5 mM, tCr_WM_= 5.7 mM). The number of deuterons and protons per molecule contributing to the detected resonance was accounted for. Details of the quantification are shown in Supplementary Figure 1. Glx_4_ concentrations were not corrected for ^2^H label loss in order to compare signal increase and decrease in mM units between ^2^H and ^1^H acquisition methods (38). Voxel-wise fractional water content of GM and WM was taken into account, by using automatic GM/WM/CSF segmentation on high resolution T_1_-weighted 3D images using the FAST algorithm (39), followed by down-sampling to MRSI grid-size using MINC tools (MINC tools, v2.0, McConnell Brain Imaging Center, Montreal, QC, Canada). For regional averaging over GM and WM, an 80% threshold was used to minimize partial volume effects.

### Statistical Analysis

Pearson correlation analysis was performed between regionally averaged 7T DMI and 3T QELT metabolite concentration time courses (Glx_4_ and Glc_6_) and between concentrations and time. Rank transformation of non-parametric tests are not recommended for small samples sizes and there is no fundamental objection of using a regular t-test(40). Therefore, paired t-test was favored over non-parametric tests to estimate differences between groups with a statistical significance threshold of *p*<0.05. To correct for multiple testing p-values were adjusted using Benjamini and Hochberg method(41). Linear fitting over time was performed for Glx_4_ signals (^2^H and ^1^H) averaged over GM and WM, while Glc_6_ signals were fitted mono exponentially. Statistical tests were performed using Python 3.10 (www.python.org, packages: scipy.stats).

### 7T ^2^H DMI vs. 3T ^1^H QELT comparison

Comparison and statistical analysis between the dynamics of Glx_4_ and Glc_6_ over time acquired using 7T ^2^H DMI (increasing concentrations) and 3T ^1^H QELT (decreasing concentrations) were performed on unsigned individual results. As the time scale of data acquisition differs between 7T ^2^H DMI and 3T ^1^H QELT experiments (differences within few min), concentration estimates (^2^H: Glx_4_, Glc_6 1_H: Glx_4_) and ratios to tCr (^1^H Glc_6_) were time matched via linear (Glx_4_) and mono-exponential (Glc_6_) interpolation using individual time constants from each participant to perform correlation analysis.

Absolute Glx_4_ increase/drop (in mM) acquired using 7T ^2^H DMI/ 3T ^1^H QELT at the end of the experiment was compared between two close time points (60 and 62 min) and identical time points (using interpolation) and results were tested for significant differences.

### Data availability statement

Data generated by postprocessing methods (i.e., metabolic maps, LCModel basis sets, script files for data handling) are available online at: (https://github.com/MRSI-HFMR-Group-Vienna/DMIvsQELT.git). Raw data files are too large to be shared publicly and are available from the corresponding author on reasonable request for research purposes only. Due to data protection policy 3D high resolution images are only available upon reasonable request from the corresponding author if approved by the requesting researcher’s local ethics committee.

## Results

### Study protocol

Initial preparation scans following oral tracer administration of deuterium labeled glucose were finished 7±2min for both 7T ^2^H DMI and 3T ^1^H QELT measurements.

### 7T ^2^H DMI

Strong positive correlation with time was observed for regionally averaged ^2^H Glx_4_ concentrations in GM (r=0.97, *p*<0.001) and WM (r=0.97, p<0.001) over all participants. After 67 min concentrations were significantly increasing to 1.36±0.16 mM (p=0.003) and 1.18±0.15 mM (p=0.002), in GM and WM, respectively. Slopes of the linear regression revealed a 14±3% faster signal (p=0.02) increase (steeper slopes) in GM (21±3 µM/min) compared to WM (19±2 µM/min) see Figure 1a.

**Figure 1:**
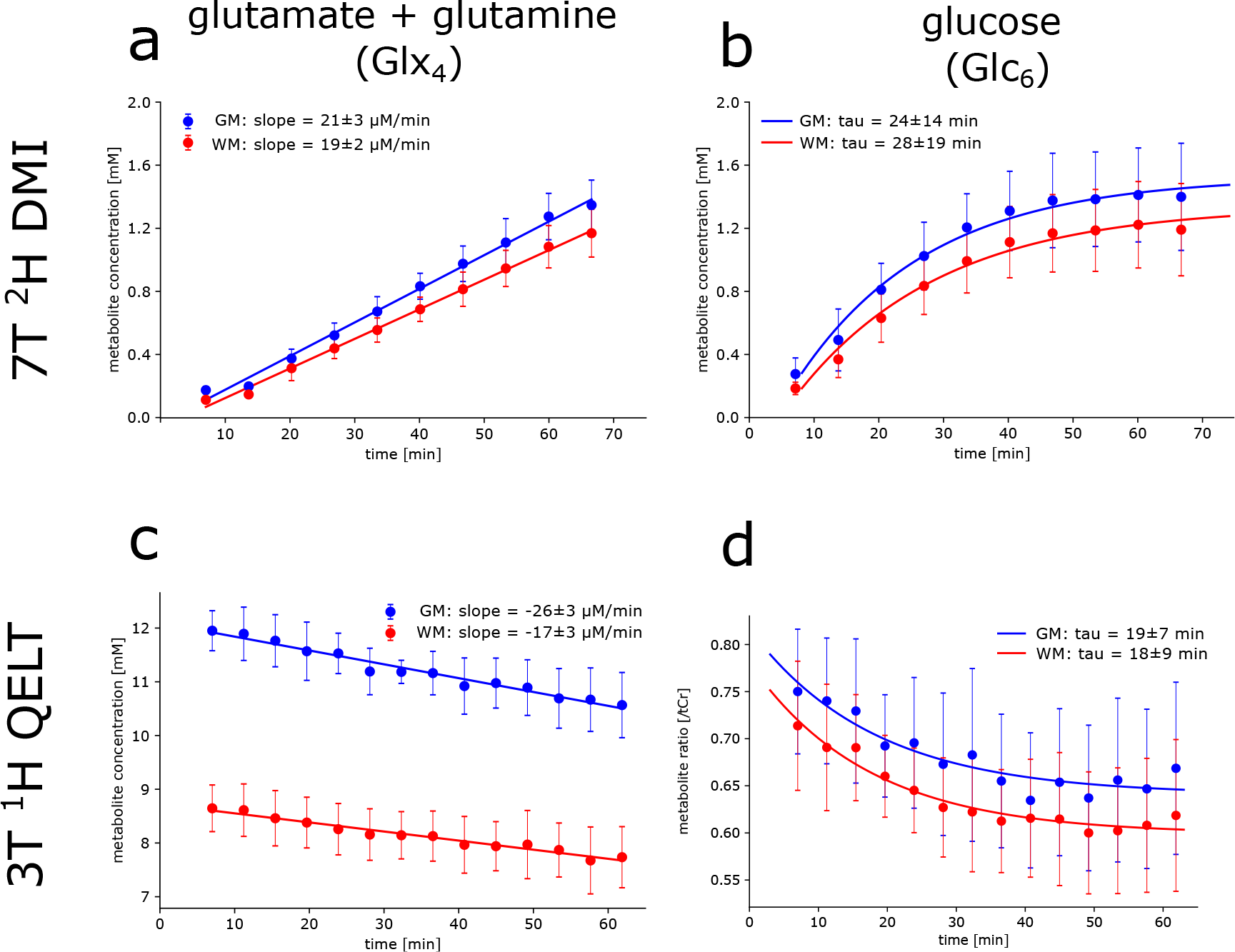
Time courses of ^2^H (top) and ^1^H (bottom) resonances from deuterium labeled (a,c) glutamate+glutamine (Glx_4_) and (b,d) glucose (Glc_6_) acquired using direct and indirect deuterium detection, i.e., 7T ^2^H DMI and 3T ^1^H QELT, respectively. Signal concentration estimates given in mM (a,b,c) and ratios to total creatine (d) were calculated voxel-wise and averaged over gray matter (GM, blue) and white matter (WM, red) dominated regions and over all participants. Concentration estimates and dynamics of increasing ^2^H Glx_4_ signals were not significantly different to decreasing ^1^H Glx_4_ acquired using DMI at 7T and QELT at clinical 3T, respectively in GM (1.29±0.15 vs. 1.38±0.26 mM, p=0.65 and 21±3 vs. 26±3 µM/min, p=0.22) and WM (red: 1.10±0.13 vs. 0.91±0.24 mM, p=0.34 and 19±2 vs. 17±3 µM/min, p=0.48). No significant differences were found between increasing ^2^H Glc_6_ and decreasing ^1^H Glc_6_ exponential time constants in GM (24±14 vs. 19±7 min, p=0.65) and WM (28±19 vs. 18±9 mM, p=0.43).

Similarly, strong positive correlation with time was observed for ^2^H Glc_6_ concentrations in GM (r=0.78, p<0.001) and WM (r=0.80, p<0.001), significantly increasing after 67 min to 1.42±0.34 mM (p=0.02) and 1.21±0.30 mM (p=0.02), respectively. Time constants of the exponential fit were not significantly different (p=0.129) between GM (24±14 min) and WM (28±19 min) see Figure 1b. Individual results of linear regression analysis for Glx_4_ concentrations and mono-exponential fitting for Glc_6_ concentrations are shown in Table 1 for all participants.

**Table 1:** Individual concentration differences, slopes of the linear regression analysis and exponential time constants for ^2^H/^1^H Glx_4_ and Glc_6_ after 60 min (7T DMI) and 62 min (3T QELT).

| Metabolite | $\Delta\text{CGM}[\text{mM}]^*$ | $\Delta\text{CWM}[\text{mM}]^*$ | slopes GM<br>[$\mu\text{M}/\text{min}$]* | slopes WM [$\mu\text{M}/\text{min}$]* |
| --- | --- | --- | --- | --- |
| Glx<sub>4</sub> |  |  |  |  |
| Participant 1 | +1.36/-0.99 | +1.20/-0.96 | +23.9/-21.8 | +21.0/-12.7 |
| Participant 2 | +1.43/-1.29 | +1.24/-0.68 | +24.1/-25.7 | +21.2/-17.4 |
| Participant 3 | +1.06/-1.72 | +0.92/-1.32 | +17.1/-30.7 | +15.1/-21.4 |
| Participant 4 | +1.17/-1.63 | +0.95/-0.93 | +18.7/-27.7 | +17.1/-17.7 |
| Participant 5 | +1.41/-1.30 | +1.17/-0.66 | +23.5/-23.0 | +19.9/-15.4 |
| Glc<sub>6</sub> |  |  |  |  |
| | $\Delta\text{CGM}[\text{mM}]$ | $\Delta\text{CWM}[\text{mM}]^*$ | tau GM [min]* | tau WM [min]* |
| Participant 1 | +1.37/ - | +1.26/ - | 13.6/27.6 | 15.6/35.7 |
| Participant 2 | +1.8/ - | +1.56/ - | 23.7/8.4 | 25.7/11.8 |
| Participant 3 | +1.03/ - | +0.89/ - | 21.9/16.0 | 25.0/12.7 |
| Participant 4 | +1.72/ - | +1.52/ - | 49.1/20.3 | 63.6/16.1 |
| Participant 5 | +1.20/ - | +0.97/ - | 9.2/24.6 | 11.1/15.1 |

Notes. - $\Delta\text{C}$ ... Concentration estimates in mM, GM ... gray matter, WM ... white matter, tau ... exponential time constant, slopes ... inclination of linear regression analysis
\*7T $^2\text{H}$ DMI/3T $^1\text{H}$ QELT

To directly visualize glucose uptake and downstream metabolite synthesis of Glx_4_ via increasing concentrations of ^2^H signals over time, 3D metabolic maps of ^2^H Glx_4_ and ^2^H Glc_6_ are shown from one representative participant for the first and last time point, i.e., 7 min and 67 min after oral administration of deuterium labeled glucose, see Figure 2a.

**Figure 2:**
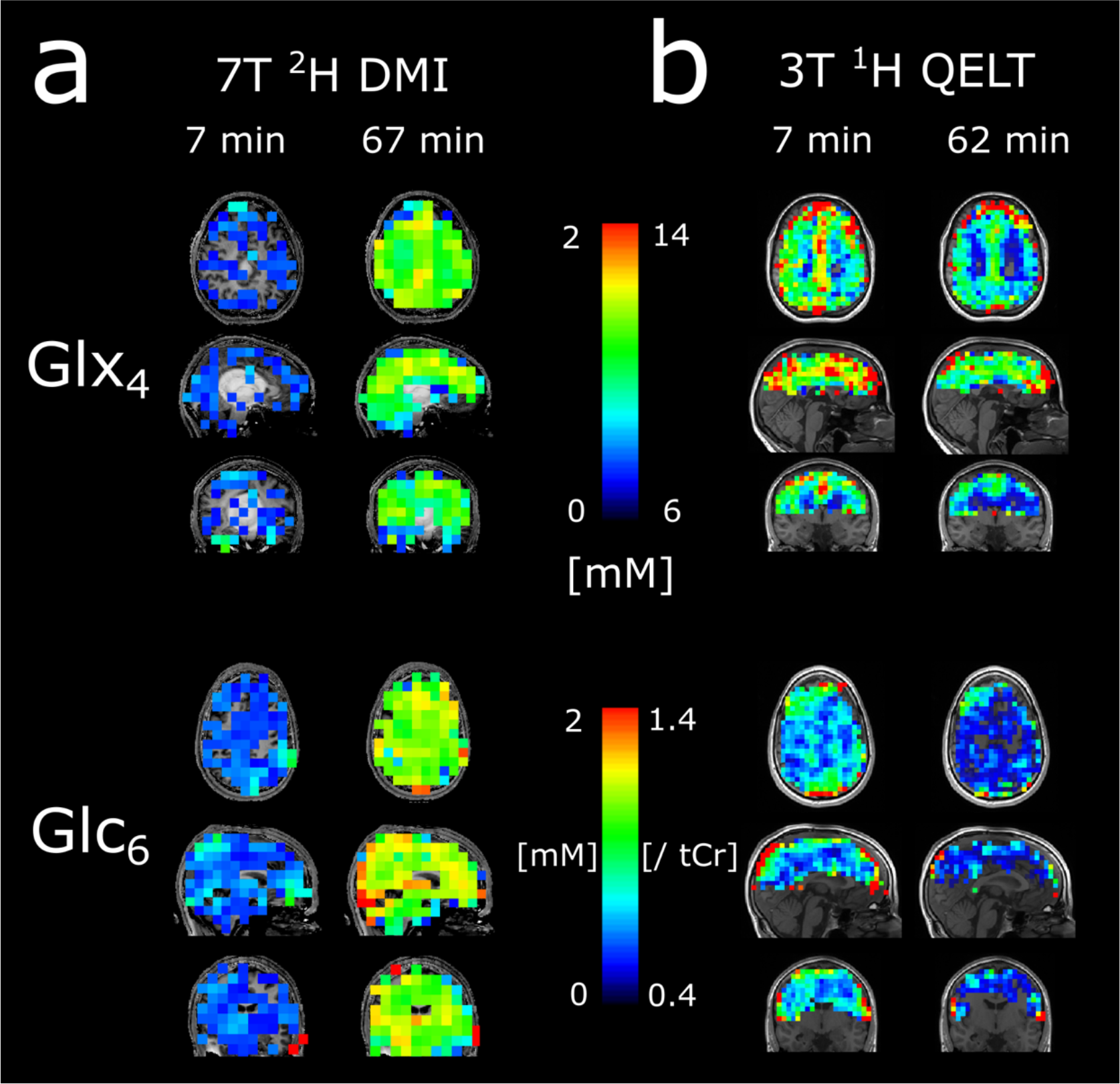
Representative 3D (glutamate+glutamine) Glx_4_ and (glucose) Glc_6_ maps of ^2^H (a) and ^1^H (b) resonances acquired 7 min and 67/62 min after deuterium labeled glucose administration. The signal intensity increase/decrease of respective ^2^H and ^1^H resonances is clearly visible.

Time courses of ^2^H Glx_4_ and ^2^H Glc_6_ metabolic maps from a representative axial slice from all participants are shown in Figure 3 and Figure 4, respectively.

**Figure 3:**
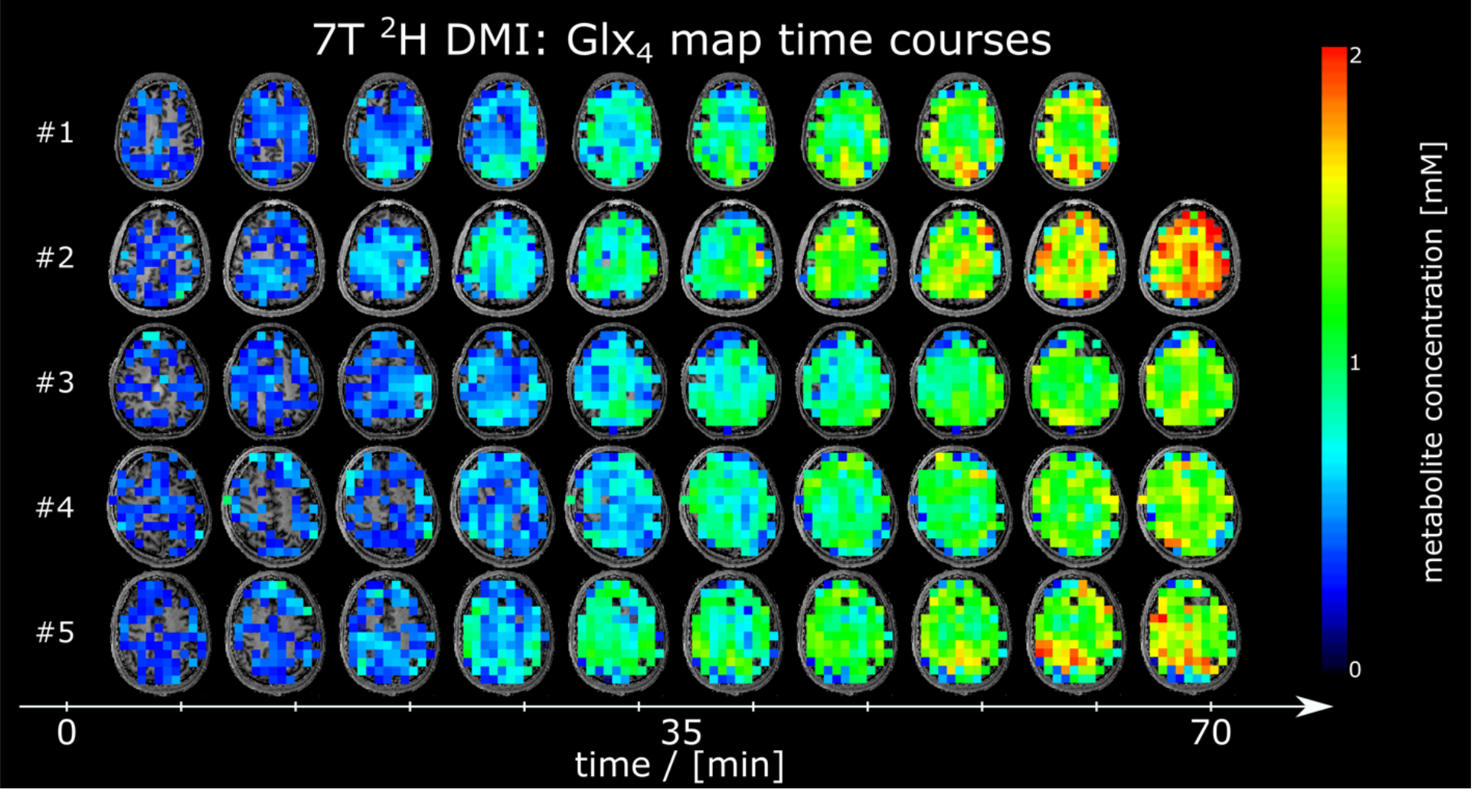
Time courses of axial ^2^H glutamate+glutamine (Glx_4_) maps given in mM from all participants, detected using deuterium metabolic imaging (DMI) at 7T. No correction of ^2^H label loss was applied.

**Figure 4:**
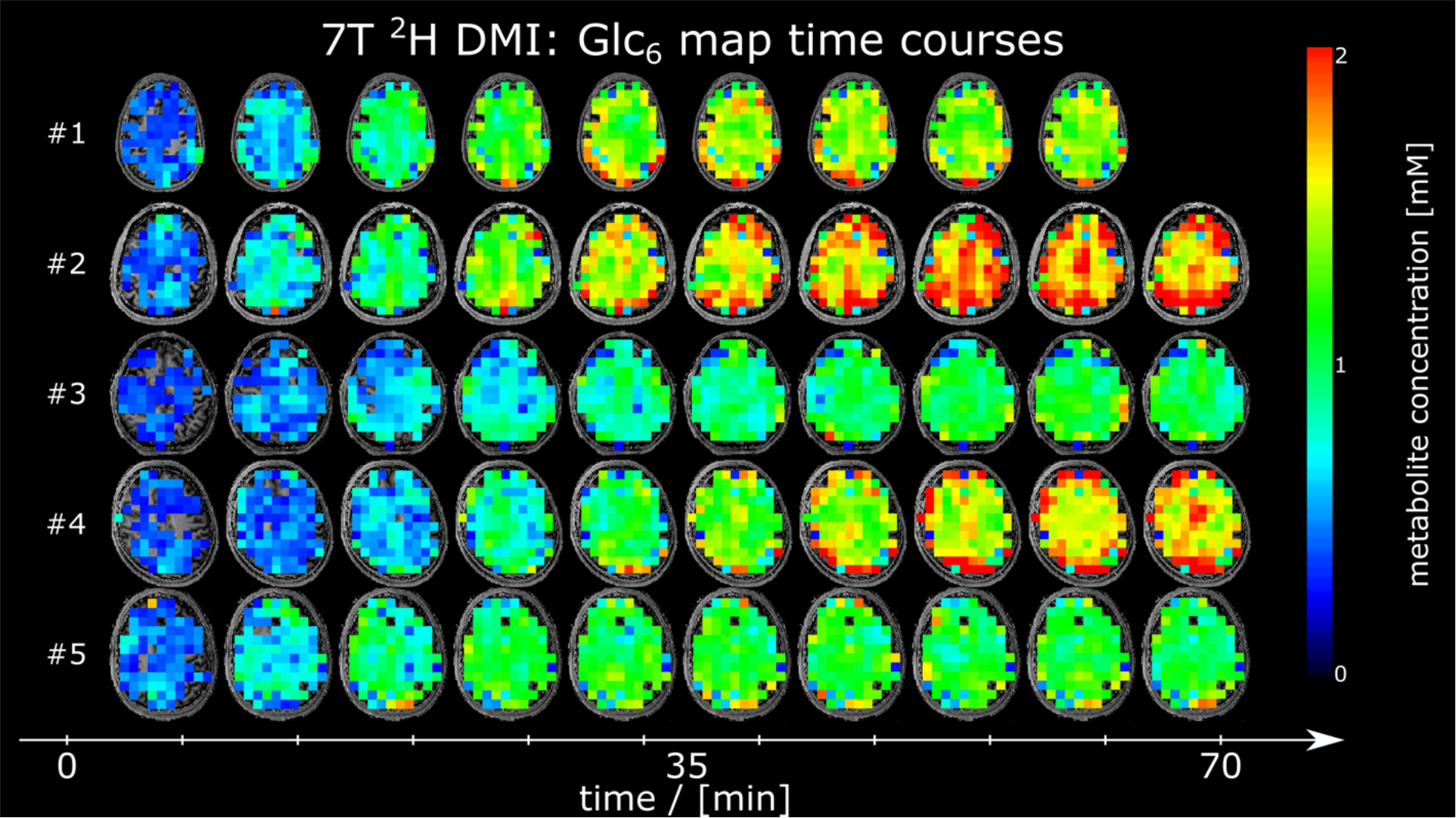
Time courses of axial ^2^H glucose (Glc_6_) maps given in mM from all participants, acquired using deuterium metabolic imaging (DMI) at 7T.

No significant changes were found between regionally averaged deuterated ^2^H water signal (GM: p=0.99, WM: p=0.52) during the first 30 min after oral uptake of deuterated glucose. Averaged time courses of deuterated water acquired using 7T ^2^H DMI for GM and WM regions and over all participants are presented in Supplementary Figure 2.

Axial SNR maps from ^2^H water and Cramer-Rao Lower Bound maps from ^2^H Glx_4_, Glc_6_ and ^2^H water are shown from one representative participant for all time points in Supplementary Figure 3

Cramer-Rao Lower bound time courses from 7T ^2^H DMI metabolites averaged over GM+WM voxels and over all participants are shown in Supplementary Figure 5a.

The standardized quality criteria used for data exclusion was fulfilled by more than 78 % for ^2^H Glx_4_ and more than 94% for ^2^H Glc_6_ quantification results in GM and WM voxels.

### 3T ^1^H QELT

Moderate to strong negative correlation with time was observed for regionally averaged ^1^H Glx_4_ concentrations in GM (r=-0.67, p<0.001) and WM (r=-0.49, p<0.001) over all participants. 62 min after oral administration of deuterium labeled glucose, ^1^H Glx_4_ concentrations decreased by -1.38±0.26 mM (p<0.001) and -0.91±0.24 mM (p=0.003) compared to the initial time point (∼7 min) in GM and WM, respectively. 54±10% faster signal (p<0.001) decrease (steeper slopes), representing faster Glx_4_ synthesis was observed in GM (-26±3 µM/min) compared to WM (-17±3 µM/min) see Figure 1c.

Weak to moderate negative correlation with time was observed for regionally averaged ^1^H Glc_6_/tCr ratios for GM (r=-0.36, p=0.002) and WM (r=-0.44, p<0.001) respectively, see Figure 1d. There was no significant difference (p=0.74) between exponential time constants of GM (19±7 min) and WM (18±9 min). Individual results of linear regression analysis of Glx_4_ concentrations and mono-exponential fitting for Glc_6_/tCr ratios are shown in Table 1 for all participants.

Figure 2b illustrates representative 3D metabolic maps of ^1^H Glx4 concentrations and ^1^H Glc_6_/tCr ratios from one participant for the first (∼7 min) and last time point (∼62 min) after tracer administration. Exchange between labeled and unlabeled molecules lead to decreasing amplitudes over time, indirectly representing uptake of ^2^H Glc_6_ and ^2^H Glx_4_ synthesis. Time courses of representative axial ^1^H Glx_4_ and ^1^H Glc_6_/tCr maps from all participants are shown in Figure 5 and Figure 6, respectively.

**Figure 5:**
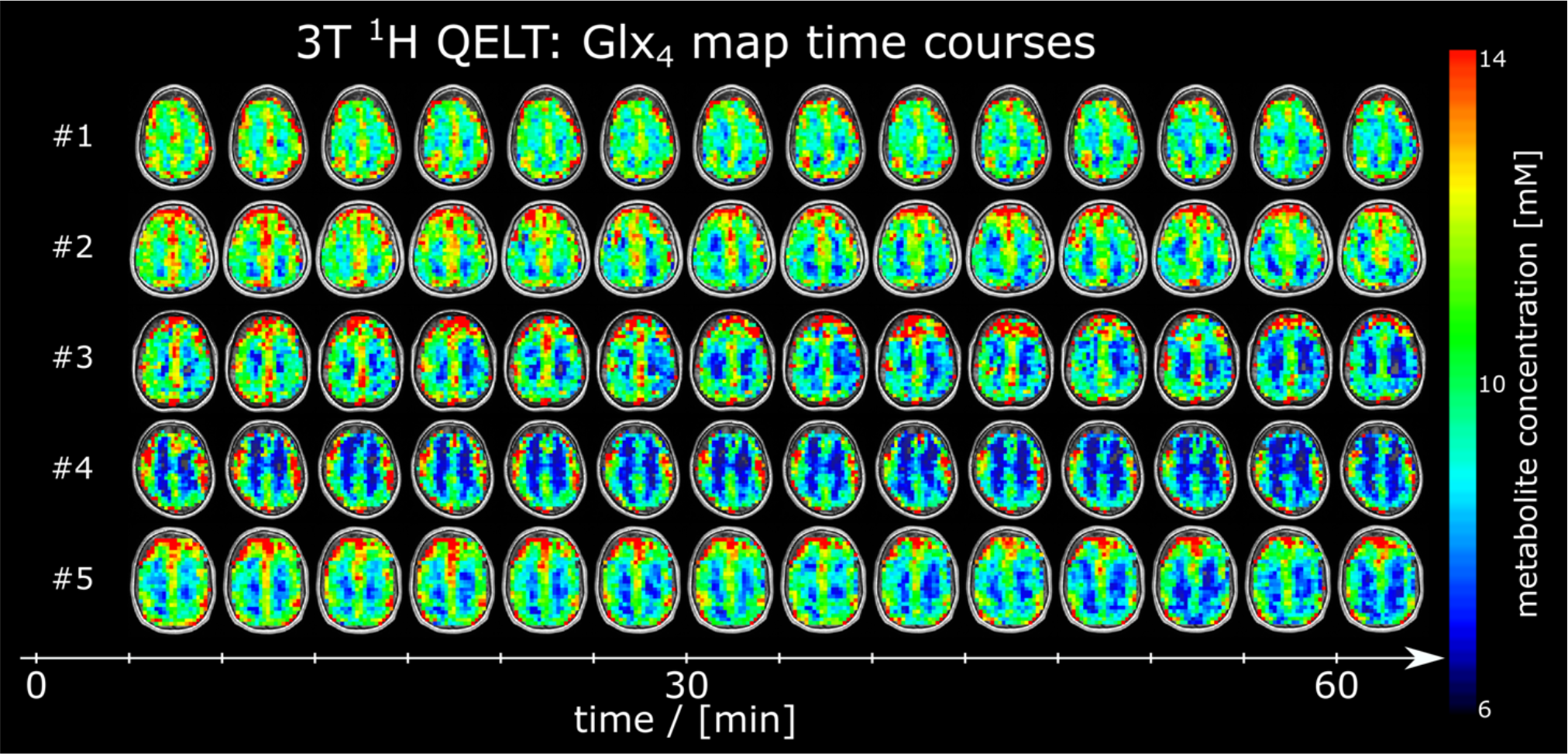
Time courses of axial ^1^H glutamate+glutamine (Glx_4_) maps given in mM from all participants, detected using quantitative exchange label turnover (QELT) at clinical 3T. No correction of ^2^H label loss was applied. The signal intensity decrease is clearly visible in all participants.

**Figure 6:**
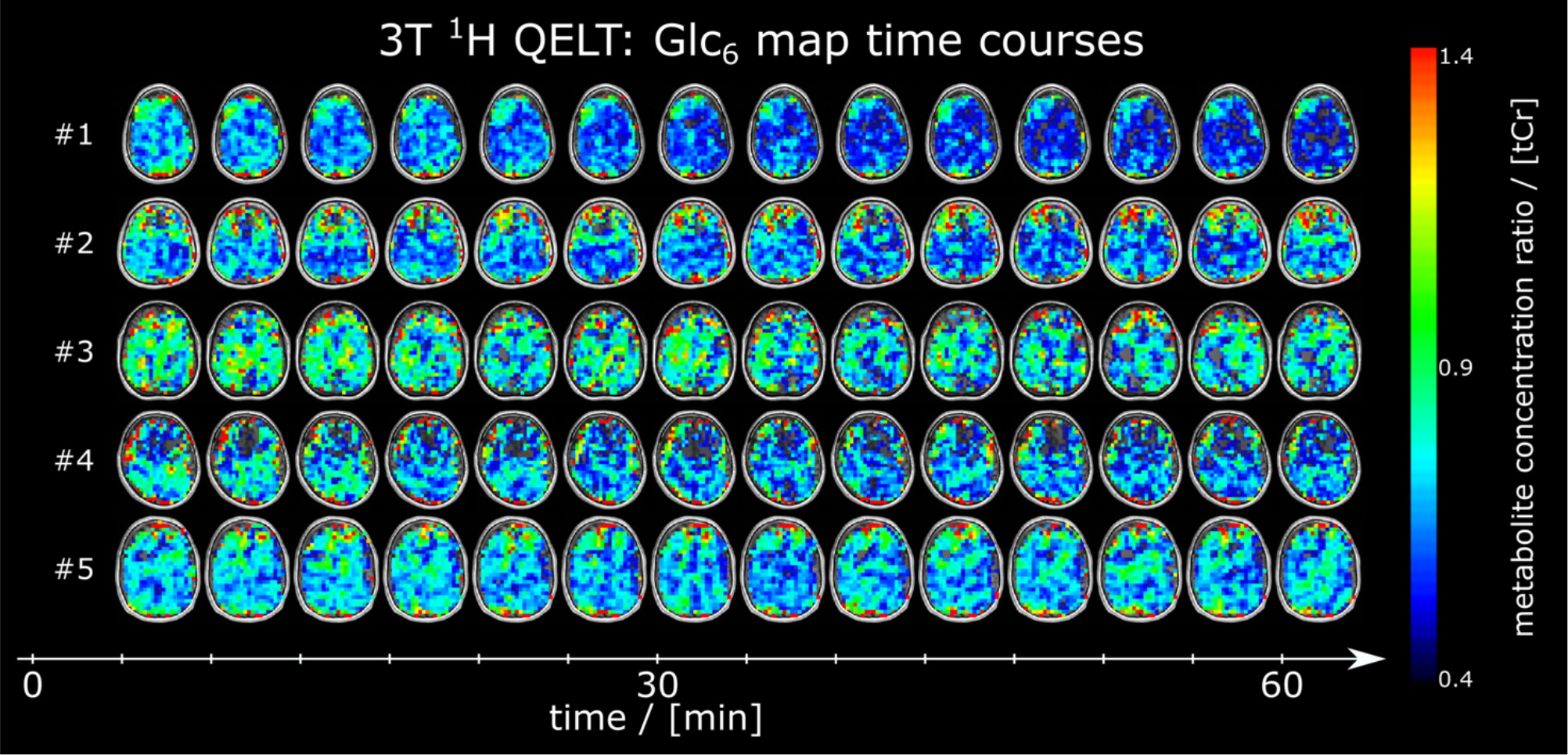
Time courses of axial ^1^H glucose (Glc_6_) maps given as ratio to total creatine (tCr) from all participants, detected using quantitative exchange label turnover (QELT) at clinical 3T.

Cramer-Rao Lower bound time courses from 3T ^1^H QELT metabolites averaged over GM+WM voxels and over all participants are shown in Supplementary Figure 5b.

Axial SNR maps from ^1^H tNAA and Cramer-Rao Lower Bound maps from all ^1^H Glx_4_, Glc_6_ and tCr metabolites are shown from one representative participant for all time points in Supplementary Figure 4. The standardized quality criteria used for data exclusion was fulfilled by more than 65 % for ^1^H Glx_4_ and more than 67% for ^1^H Glc_6_ quantification results in GM and WM voxels.

### 7T ^2^H DMI vs. 3T ^1^H QELT

60 minutes after oral consumption of deuterium labeled glucose regionally averaged ^2^H Glx_4_ concentrations were not significantly different to the decrease of ^1^H Glx_4_ concentrations after 62 min for GM (1.29±0.15 vs. 1.38±0.26 mM, p=0.65) and WM (1.10±0.13 vs. 0.91±0.24 mM, p=0.34) regions, over all participants.

Further, unsigned slopes of the linear regression analysis showed no significant differences between ^2^H and ^1^H Glx_4_ dynamics over time for GM (21±3 vs. 26±3 µM/min, p=0.22) and WM (19±2 vs. 17±3 µM/min, p=0.48) data points.

Time matched ^2^H Glx_4_ concentrations (62 min) using linear interpolation individually for each participant showed no significant differences compared to the respective ^1^H Glx_4_ concentration decrease in GM (1.31±0.15 mM, p=0.72) and WM (1.12±0.14 mM, p=0.3) dominated regions.

Correlation analysis on individual data points between averaged ^2^H and ^1^H Glx_4_ concentrations revealed a moderate negative correlation in GM (r=-0.52, p<0.001), while a weak negative correlation was observed in WM (r=-0.3, p<0.001), see Figure 7a.

**Figure 7:**
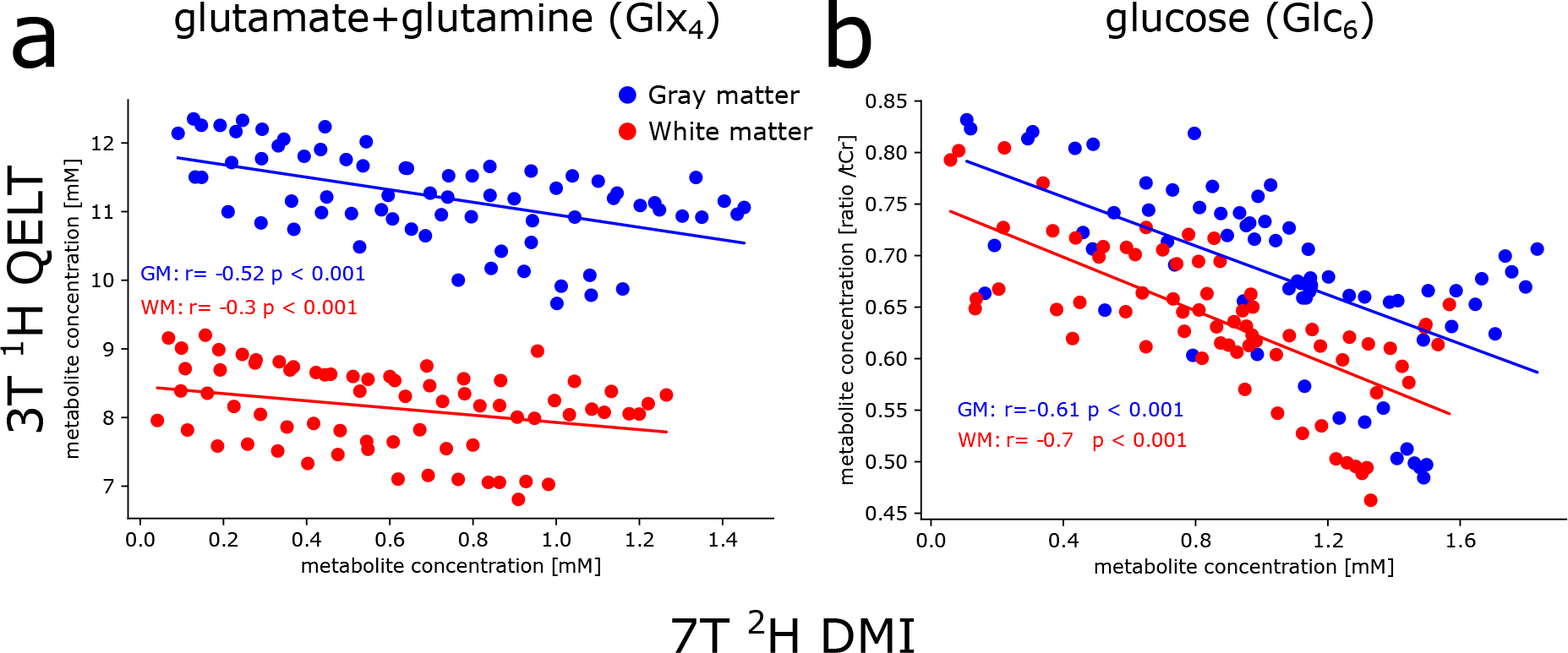
Correlation analysis between ^2^H and ^1^H resonances of Glx_4_ and Glc_6_ metabolites, averaged over gray matter (GM, blue) and white matter (WM, red) dominated regions. Data points for each participant were time matched via linear (for Glx_4_) or exponential (for Glc_6_) interpolation using individual slopes of linear regression analysis or time constants from mono-exponential fitting. Weak to moderate correlations were found between ^2^H and ^1^H signals for Glx_4_, for GM and WM regions, respectively, while strong correlation was found for Glc_6_ results.

No significant differences were observed between time constants of dynamic ^2^H Glc_6_ concentrations and ^1^H Glc_6_/tCr ratios in GM (24±14 vs. 19±7 min, p=0.65) and WM (28±19 vs. 18±9 min, p=0.43) dominated regions. A strong negative correlation was observed between individual data points of ^2^H Glc_6_ concentrations and ^1^H Glc_6_/tCr ratios in GM (r=- 0.61, p<0.001) and WM (r=-0.70, p<0.001) regions, see Figure 7b.

Representative ^2^H and ^1^H sample spectra with corresponding spectral fit of single GM and WM voxels from the first and last time points of one representative participant are presented in Figure 8. Corresponding fit output from LCModel including concentrations (given in arbitrary units) and Cramer-Rao Lower Bounds are presented in Table 2.

**Figure 8:**
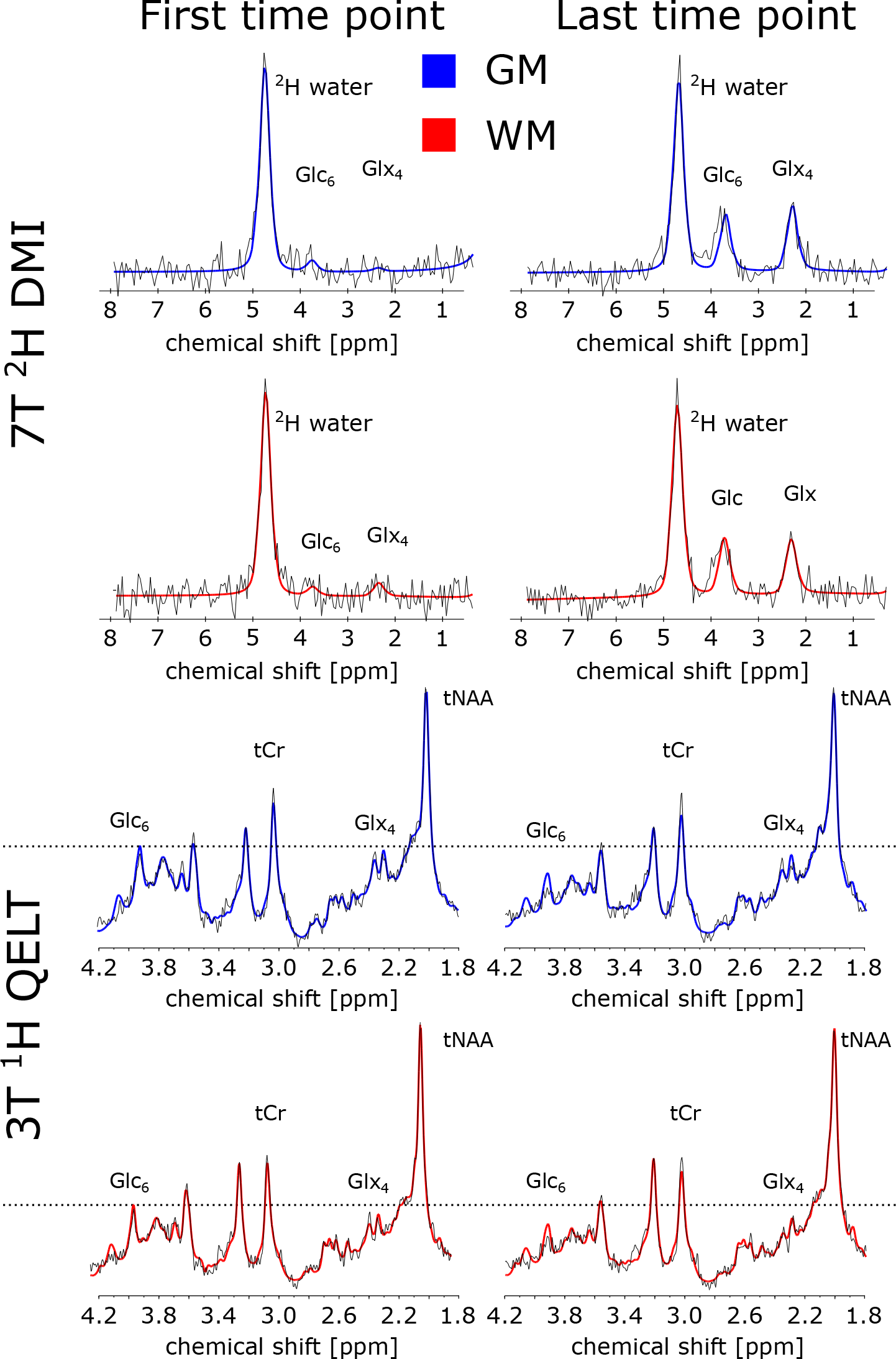
Sample spectra (black) and spectral fit results from LCModel (red) from representative gray matter (GM, blue) and white matter (WM, red) voxels acquired for the first and last time point using ^2^H DMI (a) and ^1^H QELT (b), at 7T and clinical 3T, respectively. For illustration purposes, spectra were first order corrected. Corresponding fit results of the respective voxels are shown in Table 2 for relevant metabolites, i.e., glutamate+glutamine (Glx_4_), glucose (Glc_6_), total creatine (tCr) and ^2^H water.

### Blood glucose results

Compared to baseline (0 min) glucose concentration in blood plasma increased significantly (p=0.033) from 86±7 (76–97) mg/dl to 126±25 (91–164) mg/dl at 34±2 min after oral consumption of deuterium labeled glucose. Glucose concentrations further decreased to 106±17 (86–132) mg/dl at 100±1 min on average see Supplementary Figure 6.

## Discussion

This study presents non-invasive imaging of human brain glucose metabolism with high spatial resolution, using deuterium metabolic imaging (^2^H DMI) at 7T and quantitative exchange label turnover (^1^H QELT) at clinical 3T, repetitively, in the same cohort of subjects. One hour after oral administration of deuterium labeled glucose absolute concentration estimates and the individual dynamics of accumulated deuterium labeled metabolites such as glucose (Glc_6_) and combined glutamate+glutamine (Glx_4_) were comparable between both methods and in good agreement with values reported in literature (10, 12, 14, 18).

Faster metabolic activity represented by faster deuterium labeled Glx_4_ accumulation was observed for gray matter (GM) compared to white matter (WM) dominated regions, over all participants for both methods, but was less pronounced in 7T ^2^H DMI data compared to 3T ^1^H QELT data (14% vs 54% faster), presumably due to partial volume contamination, as a result of lower spatial resolution of DMI data. Observed regional differences of metabolic activity are in the range of reported values from [^18^F]FDG-PET literature (∼33 % higher oxidative Glc consumption in GM than in WM) (42–44). Relative regional differences of TCA cycle rates (68%) reported in ^13^C-studies are higher (45, 46) compared to metabolic differences presented in this study. However, no significant differences were found for Glc_6_ dynamics between GM and WM dominated regions in this study for both 7T DMI and 3T QELT methods.

Although the nominal spatial resolution is higher in this study compared to recent DMI literature (1.95 ml vs. 2.7ml) (13, 14, 18), the point spread function is still suboptimal and still a major limitation of the currently implemented 7T DMI method to resolve fine structures of the brain. This presumably causes significant partial volume contamination, which could explain the intrasubject variability between 7T DMI and 3T QELT sessions especially in WM dominated regions and additionally smaller differences between GM and WM slopes of ^2^H Glx_4_ dynamics. Physiological variance of glucose metabolism between repeated sessions could additionally affect the results. Using spatial spectral non-cartesian sampling of the k-space (47, 48), could potentially increase the spatial resolution without prolonging scan time, but puts more stress on the gradient system due to the 6.5 times lower gyromagnetic ratio of _2_H compared to ^1^H as higher gradient amplitudes are required and additionally smaller voxel yield lower SNR (49). These challenges need to be addressed in future DMI studies.

Glx_4_ concentration estimates were not corrected for ^2^H label loss in the TCA cycle (glutamate: 38±1%, glutamine: 41±5%(38)). While it is straight forward to correct ^2^H Glx_4_ concentration estimates acquired using DMI by a fixed factor, a correction of indirectly detected ^1^H Glx_4_ concentrations can only be performed on the dynamics (Glx_4_ decrease over time). General correction of absolute ^1^H Glx_4_ estimates (including e.g., baseline measurements) would introduce a systematic error. As a correction of only ^2^H Glx_4_ results would lead to more confusion, we decided to compare results of the Glx_4_ dynamics between direct (DMI) and indirect (QELT) detection without correction of ^2^H label loss (18, 38).

To reliably detect a 10-20% decrease in signal amplitude for labeled metabolites using ^1^H QELT MRSI at 3T a high temporal stability is required, which has been tested extensively in a previous study (19) during test and re-test repeatability measurements, presenting a coefficient of variation of <2 % for stable unlabeled metabolites (i.e., tCr, tNAA, Glx_2+3_). To exclude the possibility of a systematic error causing the decrease of labeled Glx_4_ and Glc_6_ in ^1^H QELT data, control measurements using regular glucose (dextrose) were performed in another study using a similar sequence (without motion correction) at 7T (15) and were therefore, not repeated.

In general, using ^1^H MRS to detect glucose in the human brain is notoriously difficult, due to overlapping resonances between 3.5-4 ppm and inherently low SNR. However, by separating labeled from unlabeled resonances in the basis set for spectral fitting, i.e., ^1^H Glc_6_ and ^1^H Glc_1-5_, respectively, it was possible to fit ^1^H Glc_6_ (for the more abundant Glucose-Beta anomer) with a Cramer-Rao Lower Bounds threshold <50% in more than 67 % of GM and WM voxels. These results have to be interpreted carefully, however, the signal decay of ^1^H Glc_6_ (given as ratio to tCr) over time, presumably due to exchange of labeled and unlabeled molecules, could be fitted individually per subject and yielded time constants, which were not significantly different compared to ^2^H Glc_6_ results from DMI data. It has been reported, that glucose detection at 3T is more reliable compared to using ultra-high field MR scanners (≥7T) (50, 51). Therefore, we decided to include indirectly detected ^1^H Glc_6_ results using 3T ^1^H QELT in this study, which would be otherwise excluded given the conventional CRLB threshold of <20 %. Spectral fitting of the unlabeled component ^1^H Glc_1-5_ was, however, not feasible with sufficiently low CRLB. To the best of our knowledge, no relaxation times of glucose at 3T were reported in literature, and therefore only ratios to total creatine were reported for ^1^H Glc_6_ results.

One of the main limitations of ^1^H QELT MRSI is that a baseline reference scan is required, to estimate the signal drop over time, which should be assessed, ideally, before tracer administration. This study acquired the first 3D dataset on average 7±2min after oral tracer administration, where accumulation of labeled metabolites was considered negligible.

Additionally, the center of k-space yields higher contributions to signal amplitude compared to the periphery and therefore, was sampled in the beginning of each 3D readout (inside-out order of consecutive concentric ring trajectories).

In contrast to ^1^H QELT, direct deuterium detection using ^2^H DMI does not require a baseline scan to estimate absolute concentrations for a given time point, but to follow the dynamics of ^2^H Glc_6_ and ^2^H Glx_4_ over time and perform reliable exponential and linear fitting, metabolite concentrations shortly after tracer administration need to be detected, which suffer from inherently low SNR. Using relative Cramer-Rao Lower bounds to estimate the standard deviation of spectral fit results and exclude data points by applying a strict threshold is not always recommended, especially when dealing with low SNR data (52, 53). 7T DMI data during the first 20 min after tracer administration (first 3 time points) featured low SNR as a result of low label concentration. However, fitting results reflected those values and were in the expected range for ^2^H Glc_6_ and ^2^H Glx_4_, although Cramer-Rao Lower bound values were well beyond the established 20% threshold. Therefore, we decided to include fitting results into our dynamic analysis. An increased CRLB threshold of 50% was used for all following time points (>20 min) of 7T DMI data.

In one volunteer 7T DMI data was acquired only for 9 time points (∼60 min) and 16% higher nominal voxel volume before the protocol was adapted and applied for all further participants. As a result, ^2^H water concentration averaged over all subjects featured higher standard deviations for the first 9 time points, as illustrated in Supplementary Figure 2.

We are aware that the applied fitting models are not appropriate to reflect or represent real quantitative rates for neither Glx_4_ nor Glc_6_, and only serve as an approximation to illustrate and compare the dynamics between direct and indirect deuterium detection methods, i.e., 7T ^2^H DMI and 3T ^1^H QELT. Additionally, it has been previously shown in DMI literature, that deuterium labeled Glx_4_ increases approximately linearly during the first 60 min after oral tracer administration(12, 13, 18).

This study demonstrates that indirect detection of deuterium labeled compounds using ^1^H QELT MRSI is able to reproduce absolute concentration estimates of downstream glucose metabolites and the dynamics of glucose uptake compared to results from the same cohort of participants acquired using DMI at 7T. To the best of our knowledge, dynamic results of spatially resolved deuterium labeled Glc_6_ are reported for the first time using indirect detection of ^1^H QELT MRSI. Compared to DMI ^1^H QELT MRS allows for higher spatial and temporal resolution, while simultaneously detecting unlabeled metabolites and can be employed on widely available clinical 3T systems without additional hardware. This suggests significant potential for widespread application in clinical settings especially in environments with limited access to ultra-high field scanners and dedicated dual-tuned radiofrequency coils.

## Data Availability

Data generated by postprocessing methods (i.e., metabolic maps, LCModel basis sets, script files for data handling) are available online at: (https://github.com/MRSI-HFMR-Group-Vienna/DMIvsQELT.git). Raw data files are too large to be shared publicly and are available from the corresponding author on reasonable request for research purposes only. Due to data protection policy 3D high resolution images are only available upon reasonable request from the corresponding author if approved by the requesting researchers local ethics committee. Link: https://github.com/MRSI-HFMR-Group-Vienna/DMIvsQELT.git).

https://github.com/MRSI-HFMR-Group-Vienna/DMIvsQELT.git

## Acknowledgements

The financial support by the Austrian Federal Ministry for Digital and Economic Affairs, the National Foundation for Research, Technology and Development and the Christian Doppler Research Association, Austrian Science Fund and National Institute of Health is gratefully acknowledged.

## Notes

**Funders**: This work was supported by the National Institute of Health NIH R01EB031787 and the Austrian Science Fund: WEAVE I 6037 & KLI 1106 and Christian Doppler Laboratory for MR Imaging Biomarkers (BIOMAK)

### Competing Interest Statement

R. Lanzenberger received investigator-initiated research funding from Siemens Healthcare regarding clinical research using PET/MR. He is a shareholder of the start-up company BM Health GmbH since 2019.

### Clinical Trial

DRKS00013218

### Funding Statement

This work was supported by the National Institute of Health NIH R01EB031787 and the Austrian Science Fund: WEAVE I 6037 & KLI 1106 and Christian Doppler Laboratory for MR Imaging Biomarkers (BIOMAK)

### Author Declarations

I confirm that the current study was approved by the ethic commission of the Medical University of Vienna and written informed consent was obtained from all participants.

