## Supplementary Table 1 and Figure 1-6 for "Reproducibility of 3D MRSI for imaging human brain glucose metabolism using direct (^2^H) and indirect (^1^H) detection of deuterium labeled compounds at 7T and clinical 3T"

### Supplementary material

| *Minimum Reporting Standards in MR Spectroscopy checklist (according to Lin et al. NMR Biomed 2021)* | |
|---|---|
| **1. Hardware** | |
| *a. Field strength [T]* | 3T & **7T** |
| *b. Manufacturer* | Siemens |
| *c. Model (software version if available)* | Prisma Fit & **Terra dot Plus** |
| *d. RF coils: nuclei (transmit/ receive), number of channels, type, body part* | 3T: 1H RX 64 channels, head, Siemens 1H TX: body coil, **7T: 1H 1 Tx/ 32 channel RX Nova Medical, 2H/1H birdcage, Stark Contrast** |
| *e. Additional hardware* | N/A |
| **2. Acquisition** | |
| *a. Pulse sequence* | 3T: 3D FID-CRT MRSI<br>**7T: 3D FID-Phase Encoding MRSI** |
| *b. Volume of Interest (VOI) locations* | 3T: The excited 55 mm-thick slab was centered around the posterior cingulate region.<br>**7T: whole-brain** |
| *c. Nominal VOI size [$cm^3$, $mm^3$]* | 3T: 200×200×55 $mm^3$<br>**7T: (FOV) 200x200x175 $mm^3$** |
| *d. Repetition Time (TR), Echo Time (TE) [ms, s]* | 3T: TR=950 ms / 0.8 ms acquisition delay<br>**7T: TR=290 ms / 1.5 ms acquisition delay** |
| *e. Total number of Excitations or acquisitions per spectrum* | 3T: 1 average<br>**7T: 2 averages** |
| *In time series for kinetic studies* | N/A |
| *i. Number of Averaged spectra (NA) per time-point* | N/A |
| *ii. Averaging method (e.g. block-wise or moving average)* | N/A |
| *iii. Total number of spectra (acquired / in time-series)* | N/A |
| *f. Additional sequence parameters (spectral width in Hz, number of spectral points, frequency offsets); If STEAM: Mixing Time TM; If MRSI: 2D or 3D, FOV in all directions, matrix size, acceleration factors* | 3T: BW: 1325 Hz, 588 spectral points, 3D FOV: 200×200x130 $mm^3$, grid size: 32x32x21<br>**7T: BW: 500 Hz, 128 spectral points, 3D FOV: 200x200x175 $mm^3$** |
| *g. Water Suppression Method* | 3T: WET |
| *h. Shimming Method, reference peak, and thresholds for "acceptance of shim" chosen* | 3T: Standard shim + manual adjustment, water peak < 30 Hz<br>**7T: Standard shim + manual adjustment, water peak < 40 Hz** |
| *i. Triggering or motion correction method (respiratory, peripheral, cardiac triggering, incl. device used and delays)* | N/A |
| **3. Data analysis methods and outputs** | |
| *a. Analysis software* | LCModel 6.3-1 |
| *b. Processing steps deviating from quoted reference or product* | N/A |
| *c. Output measure (e.g. absolute concentration, institutional units, ratio)* | absolute concentration, ratio |
| *d. Quantification references and assumptions, fitting model assumptions* | Simulated in NMRScope-B, macromolecular background for 3T |
| **4. Data Quality** | |
| *a. Reported variables (SNR, Linewidth (with reference peaks))* | 3T: SNR was calculated using the pseudoreplica method, and linewidth as FWHM of the NAA fit<br>**7T: SNR was calculated by LCModel (peak height/residuum)** |
| *b. Data exclusion criteria* | $^1H$ 3T: SNR<15, FWHM<0.1 ppm, CRLBs > 20% for tCr and Glu+Gln ($Glx_4$), > 50% threshold for $Glc_6$<br>**$^2H$ 7T: CRLBs > 50 % for water, $Glc_6$, $Glx_4$, no CRLB threshold for first 3 time points (first 20min)** |
| *c. Quality measures of postprocessing Model fitting (e.g. CRLB, goodness of fit, SD of residual)* | CRLB |
| *d. Sample Spectrum* | See Figure 8 |

**Supplementary Table 1:**

Minimum Reporting Standards for in vivo MR Spectroscopy
Note. – Parameters 3T QELT / **7T DMI (BOLD),** CRLB = Cramér-Rao lower bounds; FID = free induction decay; CRT = concentric ring trajectory; FOV = field of view; FWHM = full-width-at-half-maximum; Glu = Glutamate; Gln = Glutamine; Glc = Glucose; tNAA = total N-acetylaspartate; SNR = signal-to-noise ratio; tCr = total creatine; VOI = volume of interest.

**$^2$H DMI:**

$$[M_{Abs}] = \frac{Amplitude_M}{Amplitude_{Water}} * \frac{f_{GM} * d_{GM} * R_{water\_GM} + f_{WM} * d_{WM} * R_{water\_WM} + f_{CSF} * d_{CSF} * R_{water\_CSF}}{(1 - f_{CSF}) * R_M} * 17.2mM * \frac{N_{water}}{N_M}$$

$$R_M = e^{-T_E/T_2} * (1 - e^{-T_R/T_1})$$

**$^1$H QELT:**

$$[M_{Abs}] = \frac{Amplitude_{glx}}{Amplitude_{tCr}} * \frac{f_{GM} * R_{tCr\_GM} + f_{WM} * R_{tCr\_WM} * WM_{factor}}{(f_{GM} * R_{glx\_GM} + f_{WM} * R_{glx\_WM})} * 7.5mM$$

$$R_M = e^{-T_E/T_2} * (1 - e^{-T_R/T_1})$$

**Supplementary Figure 1:**

Concentration estimation in mM units of $^2$H resonances detected using DMI at 7T. Metabolite amplitudes ($Amplitude_M$) were referenced to deuterated water signals ($Amplitude_{Water}$) and corrected for relaxation times $R_M$ and voxel-wise fractional water content for GM and WM and CSF tissue ($f_{GM}, f_{WM}$) with $d_{GM}$=0.78, $d_{WM}$=0.65 and $d_{CSF}$=0.97.

Concentration estimation in mM units of $^1$H resonances detected using QELT MRS at clinical 3T. Glx amplitudes ($Amplitude_{Glx}$) were referenced to total creatine signals ($Amplitude_{tCr}$) and corrected for relaxation times $R_M$ and voxel-wise fractional GM and WM content ($f_{GM}, f_{WM}$). WM content was corrected by a factor of 5.7/7.5 relative to the absolute mM concentration of tCr in GM/WM.

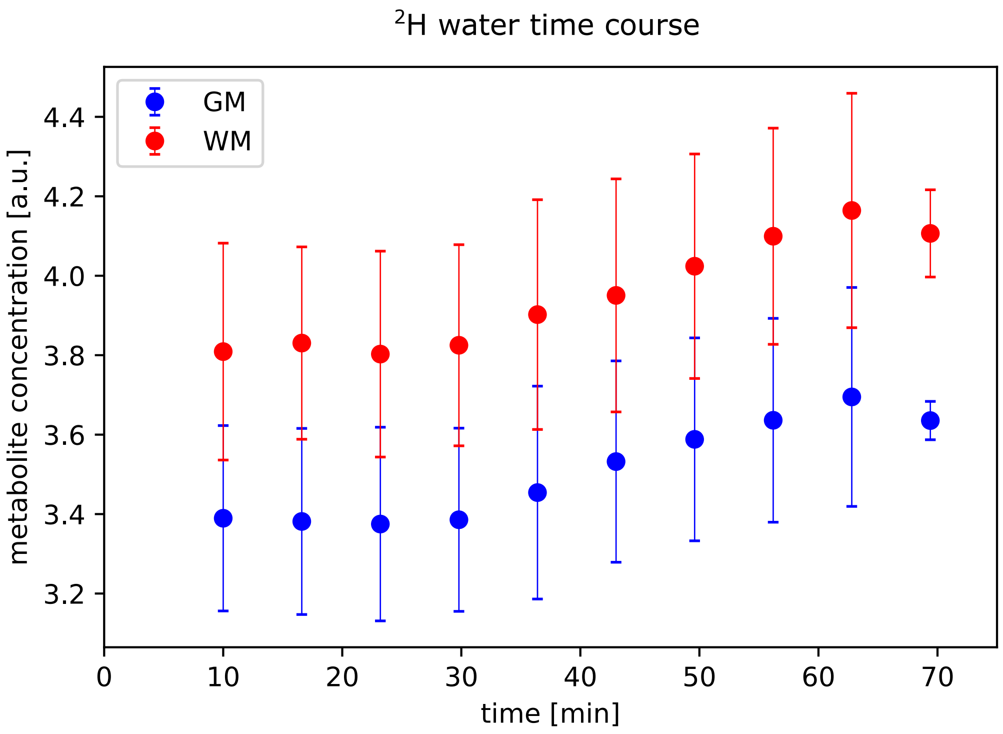


**Supplementary Figure 2:**

Averaged time courses given as (mean+-SD) of deuterated water signal resonances detected using deuterium metabolic imaging (DMI) at 7T, averaged over gray matter (GM, blue) and white matter (WM, red) dominated regions. One subject was scanned only for 9 time points and with 16% increased nominal voxel volume, which could explain the smaller standard deviation and lower mean value for the last time point.

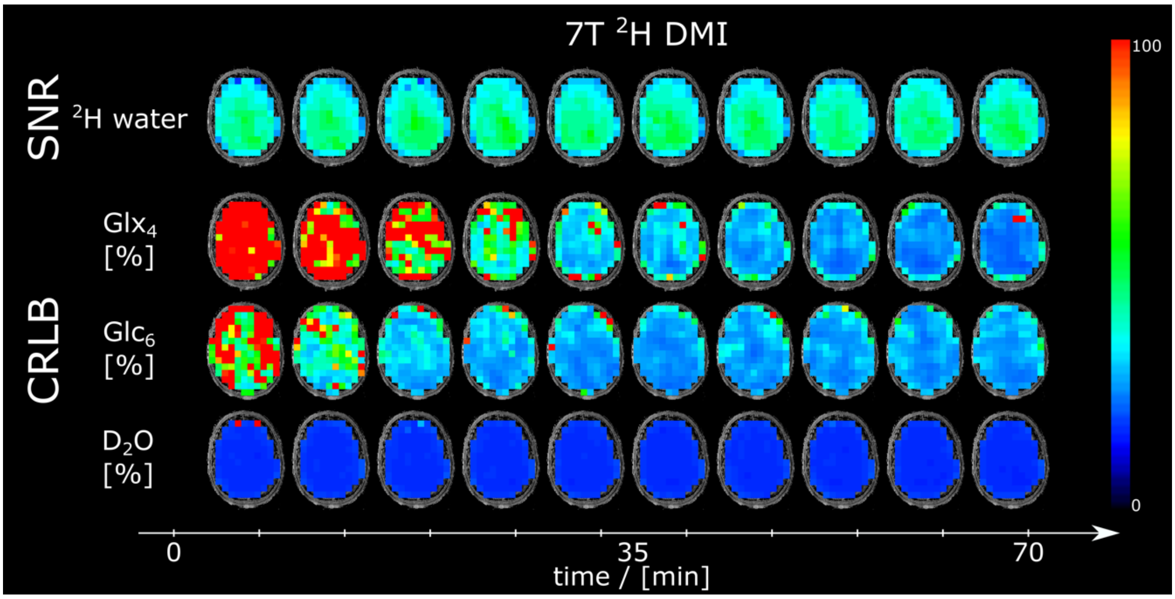


**Supplementary Figure 3:** Time courses of signal to noise ratio (SNR) maps of deuterated water and Cramer-Rao Lower Bounds maps of $^{2}H$ $Glx_4$, $Glc_6$ and deuterated water from one representative participant for all time points acquired using $^{2}H$ DMI at 7T.

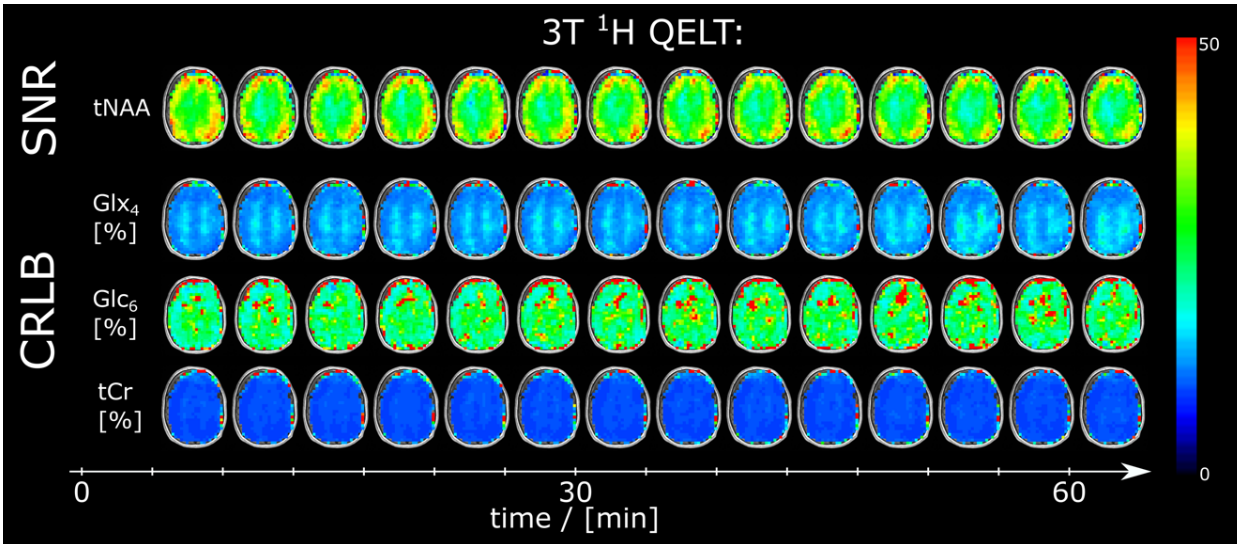


**Supplemental Figure 4:** Time courses of signal to noise ratio (SNR) maps of total N-acetyl aspartate and Cramer-Rao Lower Bounds maps of $^{1}$H $Glx_4$, $Glc_6$ and total creatine from one representative participant for all time points acquired using $^{1}$H QELT at clinical 3T.

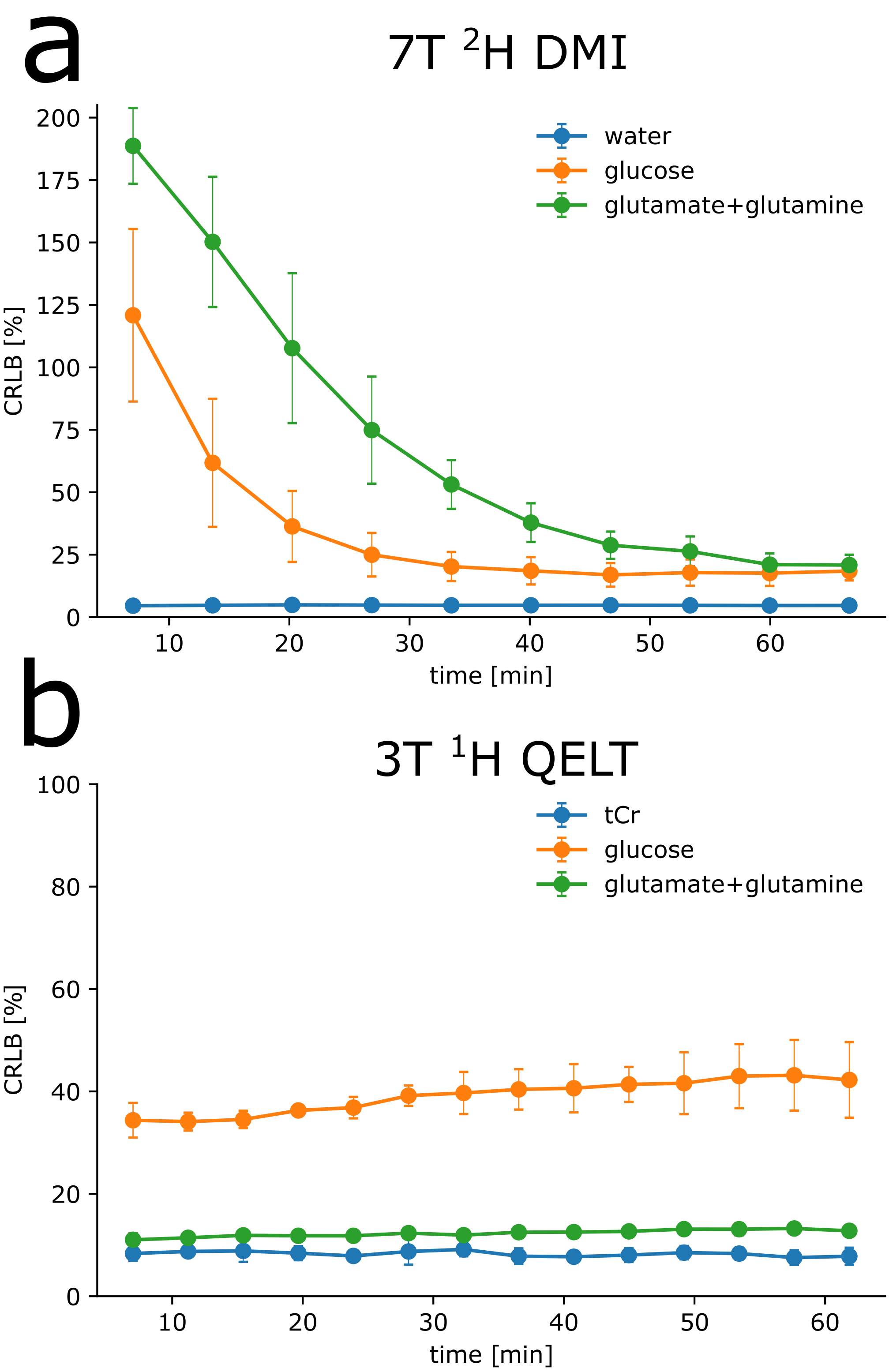


**Supplementary Figure 5:** Time courses of Cramer-Rao Lower Bounds for relevant metabolites acquired with 7T $^{2}$H DMI (a) and 3T $^{1}$H QELT (b) averaged over GM+WM voxels for each 3D dataset.

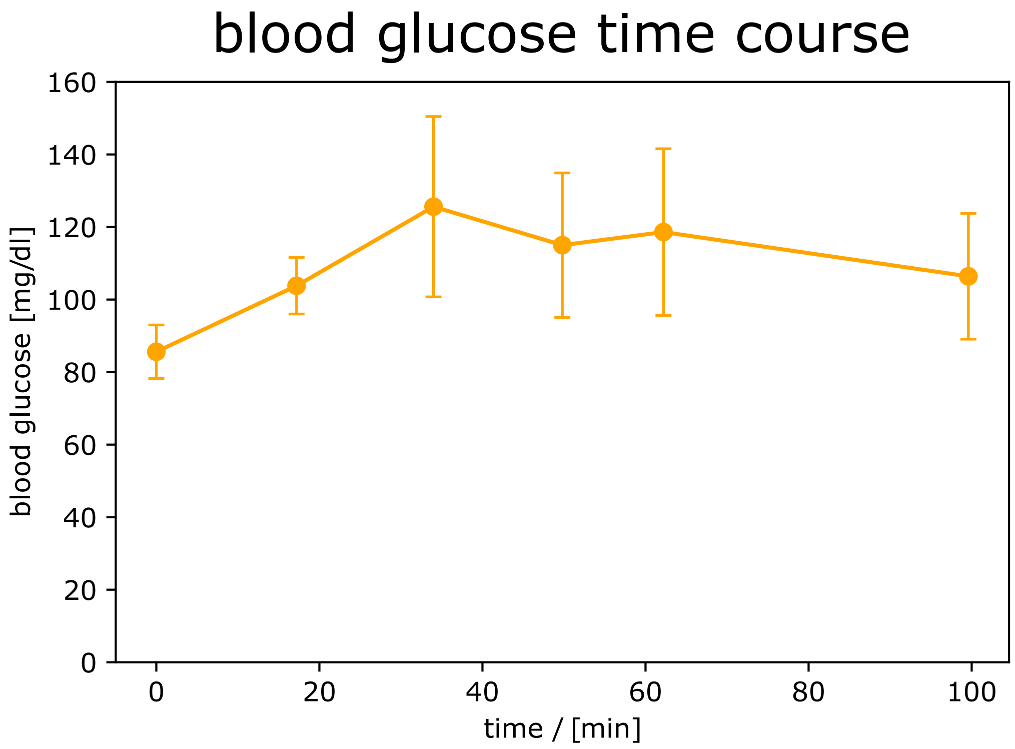


**Supplementary Figure 6:**
34±2 min after oral consumption of deuterium labeled glucose averaged blood plasma glucose concentration increased significantly (p=0.033) from 86±7 mg/dl to 126±25 mg/dl before gradually decreasing to 106±17 mg/dl at 100±1 min.
